# Individuals with and without Normal Tension Glaucoma Exhibit Comparable Performance on Tests of Cognitive Function

**DOI:** 10.1101/2020.07.17.20154468

**Authors:** Qi N. Cui, David Green, Mohit Jethi, Travis C. Porco, Jane Kuo, Todd Driver, Michael Ward, Katherine Possin, Yvonne Ou

**Affiliations:** Department of Ophthalmology, University of California San Francisco, San Francisco, California, USA; Francis I. Proctor Foundation, University of California San Francisco, San Francisco, California, USA; Department of Neurology, University of California San Francisco, San Francisco, California, USA

**Keywords:** Low Tension Glaucoma, Mental Status and Dementia Tests, Neurodegenerative Diseases, EXAMINER, CVLT-II, Executive Function, Learning and Memory

## Abstract

**Aim:** Glaucoma and dementia are both age-related neurodegenerative diseases with significant societal impact. Despite evidence suggesting an association between normal tension glaucoma (NTG) and dementia, lack of consensus remains due to conflicting reports. This cross-sectional cohort study administered a battery of neurocognitive tests targeting executive function, learning, and memory in subjects with NTG and unaffected controls to evaluate aspects of cognition impacted by dementia.

**Methods:** Fifty NTG and 50 control patients ≥ 50 years of age were recruited from the UCSF Department of Ophthalmology. Demographic data and glaucoma parameters were extracted from electronic medical records for both groups. Tests of executive function (Executive Abilities: Measures and Instruments for Neurobehavioral Evaluation and Research [EXAMINER]) and learning and memory (California Verbal Learning Test–Second Edition [CVLT-II]) were administered to both NTG and Controls. Race, handedness, best-corrected visual acuity, maximum intraocular pressure, optic nerve cup-to-disc ratio, visual field and optic nerve optical coherence tomography parameters, and a measure of general health (Charlson Comorbidity Index) were compared between NTG and Controls as well as within NTG subgroups. Multivariate linear regression was used to compare group performances on the EXAMINER battery and CVLT-II while controlling for age, sex, and years of education.

**Results:** NTG and Controls were comparable with respect to age, sex, race, education, handedness, and the Charlson Comorbidity Index (*p*>0.05 for all). Performance on the EXAMINER composite score and the CVLT-II did not differ between NTG and Controls (*p*>0.05 for both).

**Conclusions:** This is the first prospective study in which the cognitive function of subject with NTG were evaluated using a comprehensive, computerized neurocognitive battery. Subjects with NTG subjects did not perform worse than unaffected controls on tests of executive function, learning, and memory. Results do not support the hypothesis that individuals with NTG are at higher risk for cognitive dysfunction and/or dementia.

## Introduction

Glaucoma is the leading cause of irreversible blindness in the United States and is projects to exceed $7.7 billion in treatment costs in 2020^[1-3]^. Even more substantial is the monetary cost of dementia, which was in the range of $305 billion in 2020^[4]^. Both diseases increase in prevalence with age; glaucoma 3 to 16-fold from 40 to 80 years of age, and dementia 25-fold between ages 65 and 85 ^[5-8]^. Globally, as the number of older adults increases in an aging population, the number of patients with both conditions will likely follow.

In a prospective population-based cohort study, Helmer et al. 2013 found a four-fold increase in the incidence of Alzheimer’s disease (AD) among subject with open angle glaucoma (OAG) over three years.^[9]^ Interestingly, none of the subjects who later developed AD were documented as having intraocular pressures (IOP) > 21 mmHg, suggestive of an underlying diagnosis of normal tension glaucoma (NTG). This association between normal IOP and dementia in subjects with glaucoma is supported by the findings of two additional cross-sectional studies^[10,11]^. If present, a link between NTG and dementia is intriguing as it suggests an IOP-independent susceptibility to neurodegeneration that manifests in both the eye and the brain. Pathologic features associated with AD, including increased tau and decreased amyloid-beta 1-42 proteins, have been demonstrated in patients with glaucoma, while increased concentrations of apoproteins have been found in the aqueous humor of eyes with primary open angle glaucoma^[12-16]^. Structural evidence of a connection between glaucoma and neurodegenerative conditions also exists in the form of several studies demonstrating an association between decreased retinal nerve fiber layer (RNFL) and ganglion cell-inner plexiform layer thickness and cognitive decline and dementia on optical coherence tomography^[17-20]^.

Alternatively, it is possible that those diagnosed with both NTG and dementia, while exhibiting RNFL thinning and optic disc cupping that appear glaucomatous, were in fact manifesting ocular signs of a global neurodegenerative process^[21-28]^. Indeed, although a series of retrospective studies conducted in Taiwan showed an increased risk for dementia in those with OAG, several other registry-based studies found no correlation between glaucoma and dementia^[29-35]^. Still other studies attempted to answer this question by utilizing neuropsychological assessments to screen for cognitive impairments in subjects with glaucoma, yielding conflicting results^[36-38]^. For example, whereas Harrabi et al. 2015 associated a diagnosis of glaucoma with impaired performance on the Mini-Mental Status Examination blind version (MMSE-blind), Jefferis et al. 2013 demonstrated comparable performance between glaucoma subjects and controls using the same test.

To clarify whether a connection exists between NTG and neurodegenerative processes, this study administered a battery of cognitive tests to NTG and Control subjects. Tests examined executive function, learning, and memory, which are often impaired in the “mild cognitive impairment (MCI)” stage of AD and other dementing disorders^[39]^. Specifically, the Executive Abilities: Measures and Instruments for Neurobehavioral Evaluation and Research (EXAMINER) is a computer-based battery designed to provide a comprehensive evaluation of executive function^[40]^. The battery was constructed and scaled using neurological conditions such as AD, and validated via measurements of real-world executive function and correlation with dorsolateral prefrontal brain volumes^[41]^. In addition, the full-length California Verbal Learning Test–Second Edition (CVLT-II) is a widely-validated and sensitive assessment of learning and memory in various forms of dementia^[42-44]^. NTG and Control subjects demonstrated similar performance on both sets of tests. As such, the results of this study did not support the hypothesis that individuals with NTG have a higher risk for cognitive dysfunction and dementia.

## Methods

This study was conducted in accordance with the Declaration of Helsinki and the regulations of the Health Insurance Portability and Accountability Act (HIPAA). Informed consent was obtained. Institutional review board (IRB) at the University of California, San Francisco approved this study. The electronic medical records from the UCSF Department of Ophthalmology were queried on June 12, 2014 to identify patients who had at least one ophthalmology visit since January 1, 2013 and carried a diagnosis of either “low-tension open angle glaucoma” (ICD9 code 365.12) for the NTG group, or “cataracts” (ICD 9 codes 366.00-366.9) for the Control group. The EMR of these patients were then reviewed to identify patients for inclusion in the study.

Inclusion criteria were as follows: 1) age ≥ 50 years, 2) mentally capable of giving consent for participation, 3) English fluency, 4) maximum IOP (Tmax) ≤ 21 mm Hg and/or an existing diagnosis of NTG as determined by glaucoma specialists for subjects in the NTG group. Exclusion criteria were as follows: 1) best corrected visual acuity (BCVA) < 20/50 in either eye, 2) glaucoma diagnoses other than NTG, 3) diagnoses of glaucoma, glaucoma suspect, or ocular hypertension for Control subjects, and 4) an ocular history that includes exudative age-related macular degeneration, proliferative diabetic retinopathy, central retinal artery occlusion, central retinal vein occlusion, or non-glaucomatous optic neuropathy secondary to ischemic, compressive, or infiltrative causes (e.g. anterior ischemic optic neuropathy, pituitary adenoma, or intracranial hypertension).

Qualifying subjects were contacted by telephone regarding voluntary participation. 396 NTG patients were identified, of which 222 patients qualified for the study and were contacted for participation. In comparison, 715 Control patients were identified, of which 541 patients qualified for the study and were contacted for participation. The first 50 people from each group who agreed to participate were included in the study and completed all testing (Table 1). For the NTG group, glaucoma severity was determined based on the most recent Humphrey Visual Field (HVF) of the worse eye, and subjects were classified as Mild, Moderate, or Severe based upon established ICD-9 staging criteria^[45]^. The number of anti-hypertensive medications, vertical cup-to-disc ratio (CDR), as well as the most recent HVF mean deviation (MD) and the mean RNFL thickness measured by optical coherence tomography (OCT) were also collected for the NTG group.

**Table 1:** Demographic characteristics of the study population (Mean ± Standard Error).

| Group | N | Age<br>(years) | Sex<br>(M/F) | Race<br>(White/<br>Asian/<br>Hispanic/<br>Black) | Education<br>(years) | Handedness<br>(R/L) |
| --- | --- | --- | --- | --- | --- | --- |
| **NTG** | 50 | 72.5±1.2 | 14/36 | 27/17/2/4 | 16.0±0.4 | 46/4 |
| Mild | 14 | 69.6±1.9 | 1/13 | 11/2/1/0 | 16.1±0.8 | 14/0 |
| Moderate | 15 | 74.9±1.4 | 6/9 | 7/6/0/2 | 15.8±0.6 | 12/3 |
| Severe | 21 | 72.6±2.3 | 7/14 | 9/9/1/2 | 16.0±0.7 | 20/1 |
| **Control** | 50 | 70.5±1.1 | 22/28 | 32/9/6/3 | 15.7±0.4 | 46/4 |

NTG = normal tension glaucoma.

In-person testing was conducted in the UCSF ophthalmology clinic between August 2014 and March 2015 by trained examiners. Testing required 45-60 minutes to complete. At the time of testing, subjects were also queried regarding self-reported race, education, and handedness. Testing consisted of sequential administrations of: 1) the EXAMINER battery to assess executive function, and 2) the CVLT-II to assess learning and memory.

### EXAMINER

Subject were administered four tests from the EXAMINER battery: Flanker, Dot counting, N-back, and Animal fluency. A standard 15.4” Apple MacBook Pro laptop was used to administer the computerized portions of the battery, as well as to record subject response, accuracy, and reaction times. Domain scores in *Cognitive Control* (Flanker), *Working Memory* (Dot Counting and N-Back), and *Fluency* (Animal fluency), and an overall *Executive Composite* score derived from performance on all four tests, were generated for each subject using item response theory as previously described^[46]^. These scores have been shown to have good psychometric properties including no ceiling or floor effects.

The specifics of the testing paradigm are described elsewhere and are available at http://examiner.ucsf.edu^[46]^. Briefly, Flanker required subjects to observe a row of arrows pointing either to the right or to the left of the screen and to indicate the direction of the central arrow by pressing the corresponding arrow key on the keyboard. In order to respond accurately to this test of cognitive control, subjects needed to suppress all reactions to the directions of the surrounding arrows. Dot counting assessed verbal working memory and required subjects to tally, one screen at a time, the total number of dots of a specific color shown on a consecutive series of up to 7 computer screens. Subjects were then asked to recall the total number of dots from each screen in the same order in which they were presented. Similarly, N-back assessed spatial working memory by asking subjects to determine whether the location of a white square on the computer screen was the same as that of a square presented either immediately before (1-back) or two before (2-back) the current square. Finally, Animal fluency assessed verbal fluency by asking subjects to name as many animals as possible in a 60 second period.

### CVLT-II

The specifics of the testing paradigm are described elsewhere^[44]^. Briefly, the test was administered on an individual basis using standardized paper forms and began with five learning trials where a list of 16 words were read to the subject. After each trial, the subject verbally recalled as many words from the list as possible, and accurate responses were summed across these 5 trials (*Immediate Recall*). The subject was then read and recalled an interference list of 16 words. The subject then recalled the words from the first list after this short delay (*Short-Delay Free Recall*) and the after a 20-minutes long delay (*Long-Delay Free Recall*). Finally, *Long-Delay Recognition* required the subject to identify words from the first list from a list containing 32 distractor words.

### Charlson Comorbidity Index

The Charlson Comorbidity Index contains 19 medical conditions that are weighted based on the adjusted risk of mortality at one, five, and ten years for each condition^[47]^. As a substitute for a measurement of general health, an age-adjusted Charlson score representing 10-year mortality was calculated for each subject based upon ICD-9-CM and procedure codes. A higher score on the index reflects more severe comorbidity.

### Data Analysis

Multivariate linear regression models were used to compare NTG to Controls with respect to group performance on the CVLT-II and the EXAMINER battery, while controlling for age, sex, and education. A *p*-value < 0.05 was considered significant. Age, years of education, BCVA (converted to LogMAR), Tmax, CDR, HVF and OCT parameters, as well as the Charlson score, were compared between NTG subgroups (i.e. Mild, Moderate, and Severe) and between NTG and Controls using ANOVA with post-hoc Tukey HSD. Sex, race, and handedness were compared using Fisher’s Exact Test. All computations were done using R version 2.10 (R Foundation, Vienna Austria, http://www.r-project.org).

## Results

The NTG and Control groups were comparable with respect to age, sex, race, education, and handedness (*p* > 0.07; Table 1). The Charlson score was also comparable between NTG and Controls (4.44 ± 0.44 versus 3.66 ± 0.22, respectively; *p* = 0.12). BCVA was comparable between all NTG subgroups as well as between NTG and Controls (*p* > 0.51; Table 2). While Tmax was comparable between NTG subgroups (*p* > 0.12), other glaucoma-related ocular characteristics of CDR, HVF MD, and RNFL thickness were significantly different between the three NTG subgroups (*p* = 0.01, 0.01, and 0.04, respectively; Table 2).

**Table 2:**
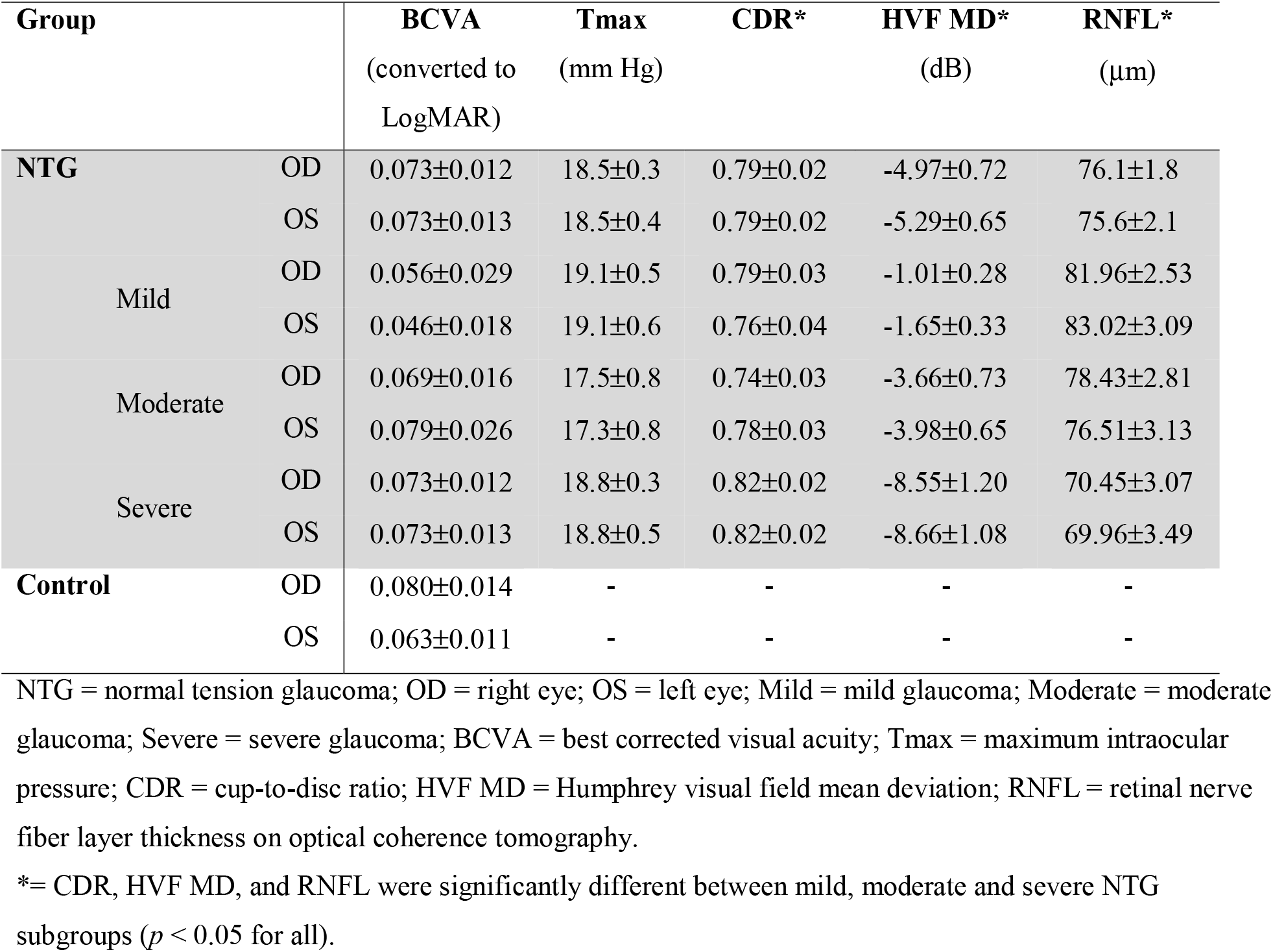
Ocular characteristics of the study population (Mean ± Standard Error).

EXAMINER *Executive Composite* and domain scores were comparable between NTG subgroups (*p* > 0.47; Figure 1 and Table 3). The *Executive Composite* and the domain scores of *Fluency* and *Working Memory* were comparable between NTG and Controls (*p* ≥ 0.06). In comparison, the domain score of *Cognitive Control* differed between NTG and Controls with the Control group performing significantly worse compared to the NTG group (*p* = 0.01). Sub-analyses comparing the NTG subgroups to Controls revealed significance only between the Moderate NTG group and Controls with respect to the domain score of *Cognitive Control (p* < 0.03).

**Table 3:**
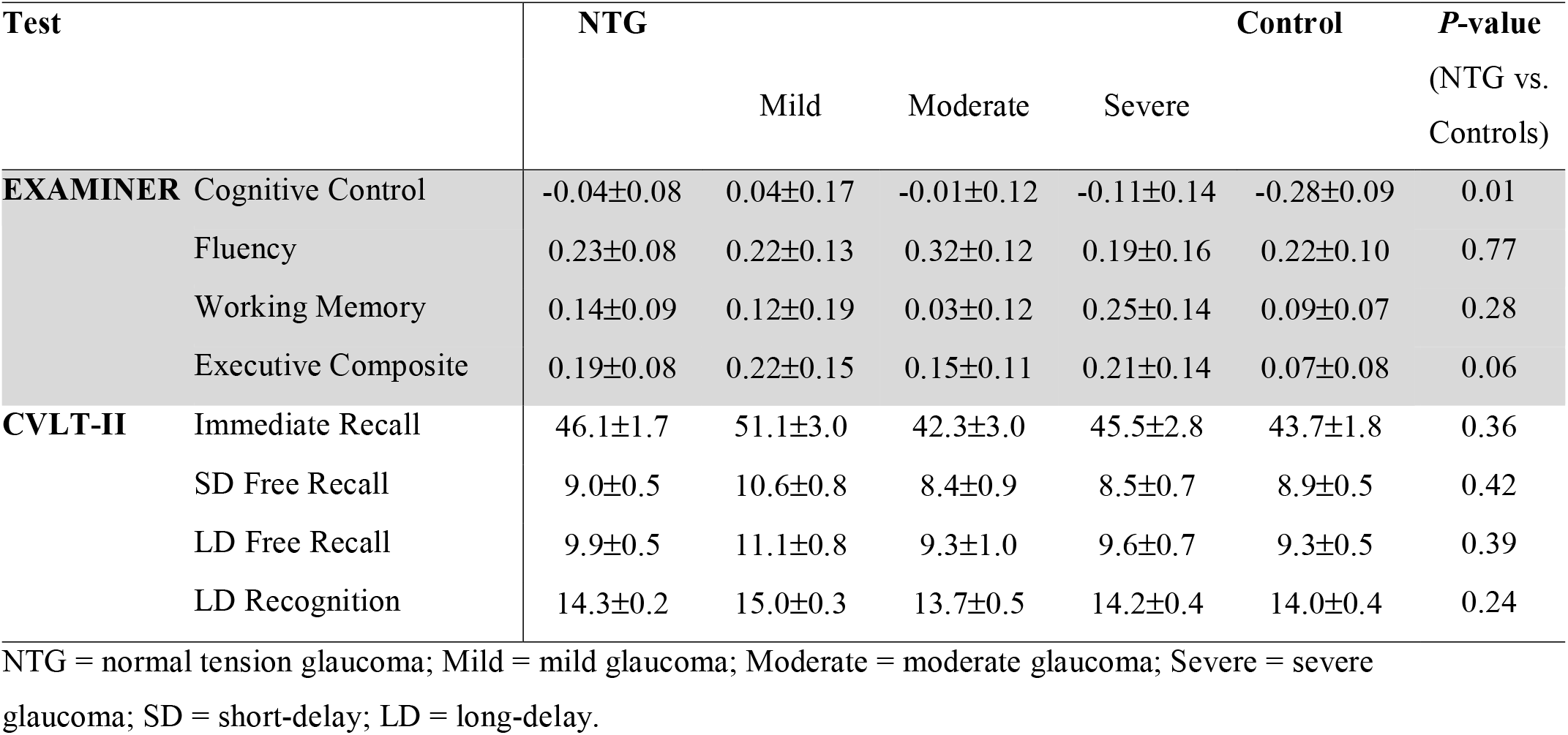
EXAMINER and CVLT-II scores for Normal Tension Glaucoma sub-groups as well as Controls (Mean ± Standard Error).

| Test | NTG |  |  |  | Control | *P*-value<br>(NTG vs.<br>Controls) |
| --- | --- | --- | --- | --- | --- | --- |
|  |  | Mild | Moderate | Severe |  |  |
| **EXAMINER** Cognitive Control | -0.04±0.08 | 0.04±0.17 | -0.01±0.12 | -0.11±0.14 | -0.28±0.09 | 0.01 |
| Fluency | 0.23±0.08 | 0.22±0.13 | 0.32±0.12 | 0.19±0.16 | 0.22±0.10 | 0.77 |
| Working Memory | 0.14±0.09 | 0.12±0.19 | 0.03±0.12 | 0.25±0.14 | 0.09±0.07 | 0.28 |
| Executive Composite | 0.19±0.08 | 0.22±0.15 | 0.15±0.11 | 0.21±0.14 | 0.07±0.08 | 0.06 |
| **CVLT-II** Immediate Recall | 46.1±1.7 | 51.1±3.0 | 42.3±3.0 | 45.5±2.8 | 43.7±1.8 | 0.36 |
| SD Free Recall | 9.0±0.5 | 10.6±0.8 | 8.4±0.9 | 8.5±0.7 | 8.9±0.5 | 0.42 |
| LD Free Recall | 9.9±0.5 | 11.1±0.8 | 9.3±1.0 | 9.6±0.7 | 9.3±0.5 | 0.39 |
| LD Recognition | 14.3±0.2 | 15.0±0.3 | 13.7±0.5 | 14.2±0.4 | 14.0±0.4 | 0.24 |

NTG = normal tension glaucoma; Mild = mild glaucoma; Moderate = moderate glaucoma; Severe = severe glaucoma; SD = short-delay; LD = long-delay.

**Figure 1:**
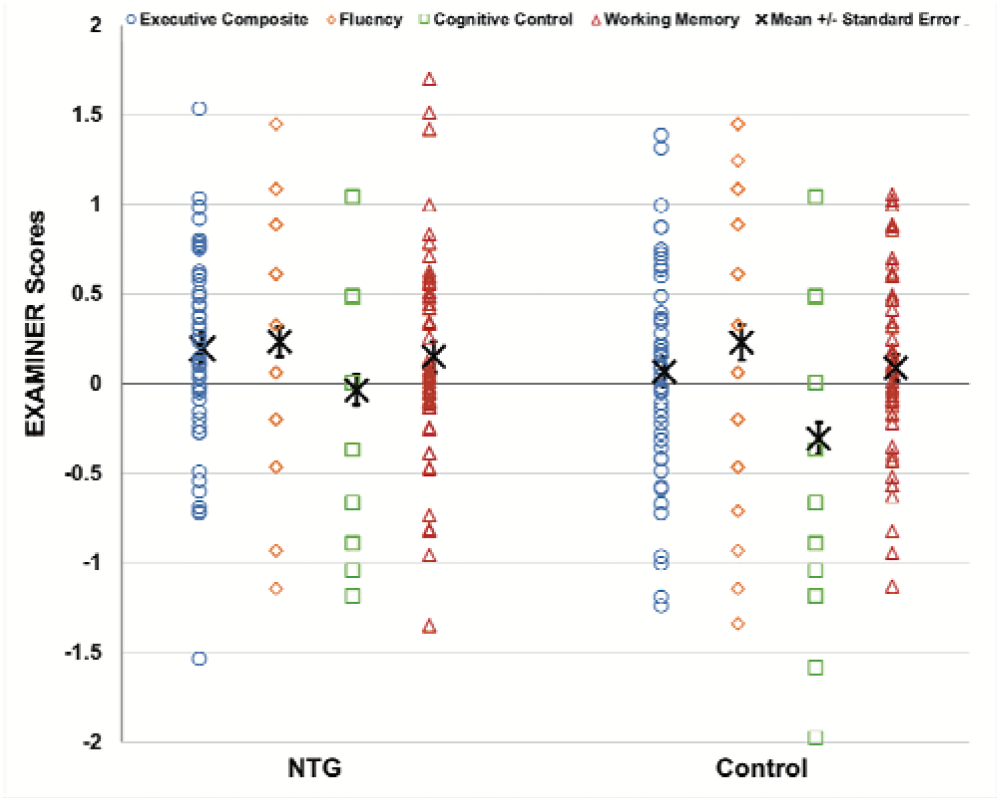
EXAMINER Executive Composite and domain scores (e.g. Fluency, Cognitive Control, and Working Memory) for Normal Tension Glaucoma and Control subjects. Error bars indicate Standard Error. NTG = normal tension glaucoma.

All CVLT-II scores were comparable between NTG subgroups as well as between NTG and Controls (*p*≥0.24 for all; Figure 2 and Table 3). A power calculation was also conducted for the EXAMINER Executive Composite and CVLT-II LD Recognition scores. For both measures, 50 cases and 50 controls provided 80% power to detect a moderate effect size of 0.57.

**Figure 2:**
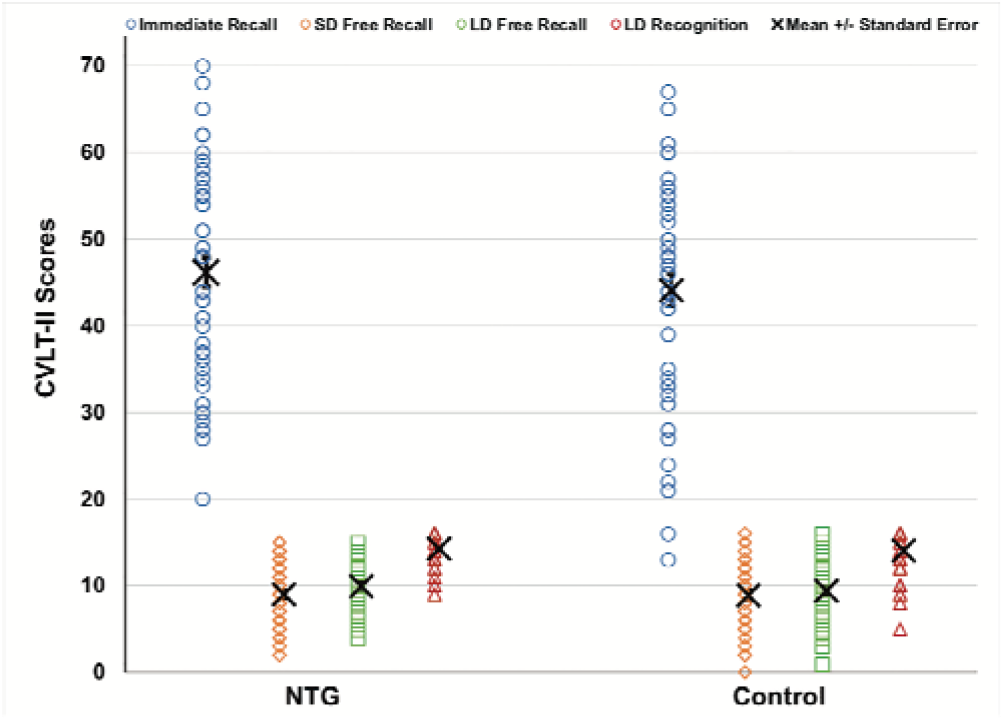
CVLT-II scores for Normal Tension Glaucoma and Control subjects. Error bars indicate Standard Error. NTG = normal tension glaucoma; SD = short-delay; LD = long-delay.

## Discussion

This is the first prospective study in which the cognitive function of subjects with NTG were evaluated using a comprehensive battery of neurocognitive tests designed to assess executive function, memory, and learning. The NTG and Control groups were well-matched with respect to age, sex, race, education, and handedness as well as the Charlson score, which served as a surrogate measure of general health. The EXAMINER *Executive Composite* score was similar between groups, as were performances on tests designed to target verbal fluency and working memory. Likewise, no difference was observed between NTG and Controls with respect to memory and learning as assessed via the CVLT-II. Testing also did not demonstrate any correlation between glaucoma severity and cognitive function.

Interestingly, the Control group performed significantly worse than the NTG group with respect to one executive function domain, *Cognitive Control*. This domain was measured by the Flanker test, which is arguably the least challenging test in the EXAMINER battery employed for this study, as it only requires correctly indicating the direction of the middle arrow. It is therefore somewhat surprising that the Controls did worse on this test compared to the NTGs while performance on all other tests were comparable between groups. One possible explanation may be a difference in the level of motivation. While many subjects in the NTG group were aware of the connection between glaucoma and dementia, the same cannot be said of the Control group. This difference in emotional investment and interest may have resulted in a difference in attention and performance between groups, which was most noticeable during Flanker testing.

Despite the outcomes of the Flanker test, taken as a whole, we found no evidence for an association between deficits in executive function, learning, and memory, and NTG. Importantly, this study was sufficiently powered to detect moderate effect sizes in our primary outcomes. The results of this study are supported by a recent publication showing similar neurocognitive function as measured by the Montreal Cognitive Assessment in open angle glaucoma patients compared to controls^[48]^. The results of this study do not support the hypothesis that glaucoma patients have worse cognitive function, which are typically evident in MCI or dementia, and thus increased risks for dementia. Our results are also in line with findings from multiple registry-based studies^[32-35]^.

Controversy exists as to whether low-vision directly impacts cognitive function. Jefferis et al. 2013 associated glaucoma with poor performance on the MMSE but not on the less visually-demanding MMSE-blind. In contrast, Harrabi et al. 2015 demonstrated lower performance on the MMSE-blind in those with vision loss due to age-related macular degeneration, Fuchs’ corneal dystrophy, and glaucoma compared to sighted individuals. To preclude any potential confounding effect of visual impairment, all participants in this study had BCVA ≥ 20/50 in both eyes.

The Helmer et al. 2013 study, which found a four-fold increase in the incidence of AD among OAG subjects, may have been affected by methodological issues in subject identification. The OAG cohort was identified based upon non-mydriatic, non-stereo color disc photos. While the diagnosis was confirmed through an ophthalmology exam, the contribution of optic nerve head imaging, and to a lesser extent, visual field testing, in establishing the OAG diagnosis was unclear. Due to this categorization ambiguity, the possibility that those OAG patients who went on to an AD diagnosis actually had dementia-associated RNFL thinning and/or optic nerve atrophy cannot be excluded. RNFL thinning and optic nerve atrophy have been demonstrated in the eyes of those with AD and MCI, and given that RNFL thinning in AD and MCI are often most prominent in the superior quadrant, may well mimic glaucomatous optic neuropathy in appearance^[21-28]^. Similar subject identification issues exist in the series of population-based studies out of Taiwan, which in combination suggested a higher risk for AD in those with OAG^[29-31]^. In those studies, the glaucoma cohort was identified based upon ICD-9-CM codes and glaucoma treatments without the benefit of additional chart review or ophthalmic examination. The criteria by which these subjects received the OAG diagnosis were not reported.

Strengths of our study include the robustness of the cognitive tests employed. The EXAMINER battery is a comprehensive and well-validated assessment of executive function. Likewise, the CVLT-II is a well-validated and widely-utilized test for learning and memory. Used in combination, these two tests provided coverage for multiple elements of cognition known to be affected in MCI and early dementia. Furthermore, unlike other studies which are based on registry information alone, all subjects in this study were: 1) directly evaluated by ophthalmologists in a glaucoma specialty ophthalmology clinic, and 2) underwent chart review by glaucoma specialists prior to enrollment to ensure correct diagnoses of NTG for the glaucoma group or lack thereof for the Control group. A possible limitation of this study is the fact that examiners were not blinded to NTG or Control status, which could have introduced bias in testing administration.

In summary, this study evaluated cognitive impairment in a population of NTG subjects utilizing a battery of tests designed to assess executive function and memory. Results do not support an increased risk for cognitive dysfunction in subjects with NTG compared to Controls without glaucoma. Future studies with a larger cohort and prospective follow-up involving repeat testing are needed to elucidate the relationship between glaucoma and cognitive dysfunction before determinations about the presence or absence of a true connection can be made.

## Data Availability

The authors confirm that the data supporting the findings of this study are available within the article [and/or]can be shared by corresponding author upon request.

## Foundation

The authors would like to acknowledge Drs. Cynthia Chiu, Ying Han, Shan Lin, Saras Ramanathan, and Robert Stamper for their contributions to patient recruitment, as well as Dr. Ari Green for assisting with project conception. The authors would also like to acknowledge the NIH-NEI EY002162 - Core Grant for Vision Research and the Research to Prevent Blindness Unrestricted Grant to the UCSF Department of Ophthalmology.

## References

1. Quigley HA, Broman AT. The number of people with glaucoma worldwide in 2010 and 2020. Br J Ophthalmol. 2006;90(3):262–267.

2. Varma R, Lee PP, Goldberg I, Kotak S. An assessment of the health and economic burdens of glaucoma. Am J Ophthalmol. 2011;152(4):515–522.

3. America PB. The future of vision: forecasting the prevalence and costs of vision problems. Prevent Blindness America. http://forecasting.preventblindness.org/disease-projections/visual-impairment-and-blindness. Published 2014. Accessed April 20, 2017.

4. Alzheimer’s A. 2020 Alzheimer’s disease facts and figures. Alzheimers Dement. 2020.

5. Tielsch JM, Sommer A, Katz J, Royall RM, Quigley HA, Javitt J. Racial variations in the prevalence of primary open-angle glaucoma. The Baltimore Eye Survey. JAMA. 1991;266(3):369–374.

6. Klein BE, Klein R, Sponsel WE, et al. Prevalence of glaucoma. The Beaver Dam Eye Study. Ophthalmology. 1992;99(10):1499–1504.

7. Varma R, Ying-Lai M, Francis BA, et al. Prevalence of open-angle glaucoma and ocular hypertension in Latinos: the Los Angeles Latino Eye Study. Ophthalmology. 2004;111(8):1439–1448.

8. Ferri CP, Prince M, Brayne C, et al. Global prevalence of dementia: a Delphi consensus study. Lancet. 2005;366(9503):2112–2117.

9. Helmer C, Malet F, Rougier MB, et al. Is there a link between open-angle glaucoma and dementia? The Three-City-Alienor cohort. Ann Neurol. 2013;74(2):171–179.

10. Bayer AU, Keller ON, Ferrari F, Maag KP. Association of glaucoma with neurodegenerative diseases with apoptotic cell death: Alzheimer’s disease and Parkinson’s disease. Am J Ophthalmol. 2002;133(1):135–137.

11. Tamura H, Kawakami H, Kanamoto T, et al. High frequency of open-angle glaucoma in Japanese patients with Alzheimer’s disease. J Neurol Sci. 2006;246(1-2):79–83.

12. Inoue T, Kawaji T, Tanihara H. Elevated levels of multiple biomarkers of Alzheimer’s disease in the aqueous humor of eyes with open-angle glaucoma. Invest Ophthalmol Vis Sci. 2013;54(8):5353–5358.

13. Gupta N, Fong J, Ang LC, Yucel YH. Retinal tau pathology in human glaucomas. Can J Ophthalmol. 2008;43(1):53–60.

14. Nucci C, Martucci A, Martorana A, Sancesario GM, Cerulli L. Glaucoma progression associated with altered cerebral spinal fluid levels of amyloid beta and tau proteins. Clin Exp Ophthalmol. 2011;39(3):279–281.

15. Yoneda S, Hara H, Hirata A, Fukushima M, Inomata Y, Tanihara H. Vitreous fluid levels of beta-amyloid((1-42)) and tau in patients with retinal diseases. Jpn J Ophthalmol. 2005;49(2):106–108.

16. Chiasseu M, Cueva Vargas JL, Destroismaisons L, Vande Velde C, Leclerc N, Di Polo A. Tau Accumulation, Altered Phosphorylation, and Missorting Promote Neurodegeneration in Glaucoma. J Neurosci. 2016;36(21):5785–5798.

17. Liu YL, Hsieh YT, Chen TF, et al. Retinal ganglion cell-inner plexiform layer thickness is nonlinearly associated with cognitive impairment in the community-dwelling elderly. Alzheimers Dement (Amst). 2019;11:19–27.

18. Ko F, Muthy ZA, Gallacher J, et al. Association of Retinal Nerve Fiber Layer Thinning With Current and Future Cognitive Decline: A Study Using Optical Coherence Tomography. JAMA Neurol. 2018;75(10):1198–1205.

19. Cunha LP, Lopes LC, Costa-Cunha LV, et al. Macular Thickness Measurements with Frequency Domain-OCT for Quantification of Retinal Neural Loss and its Correlation with Cognitive Impairment in Alzheimer’s Disease. PLoS One. 2016;11(4):e0153830.

20. Liu S, Ong YT, Hilal S, et al. The Association Between Retinal Neuronal Layer and Brain Structure is Disrupted in Patients with Cognitive Impairment and Alzheimer’s Disease. J Alzheimers Dis. 2016;54(2):585–595.

21. Blanks JC, Torigoe Y, Hinton DR, Blanks RH. Retinal pathology in Alzheimer’s disease. I. Ganglion cell loss in foveal/parafoveal retina. Neurobiol Aging. 1996;17(3):377–384.

22. Hinton DR, Sadun AA, Blanks JC, Miller CA. Optic-nerve degeneration in Alzheimer’s disease. N Engl J Med. 1986;315(8):485–487.

23. den Haan J, Verbraak FD, Visser PJ, Bouwman FH. Retinal thickness in Alzheimer’s disease: A systematic review and meta-analysis. Alzheimers Dement (Amst). 2017;6:162–170.

24. Coppola G, Di Renzo A, Ziccardi L, et al. Optical Coherence Tomography in Alzheimer’s Disease: A Meta-Analysis. PLoS One. 2015;10(8):e0134750.

25. Polo V, Garcia-Martin E, Bambo MP, et al. Reliability and validity of Cirrus and Spectralis optical coherence tomography for detecting retinal atrophy in Alzheimer’s disease. Eye (Lond). 2014;28(6):680–690.

26. Bayhan HA, Aslan Bayhan S, Celikbilek A, Tanik N, Gurdal C. Evaluation of the chorioretinal thickness changes in Alzheimer’s disease using spectral-domain optical coherence tomography. Clin Exp Ophthalmol. 2015;43(2):145–151.

27. Marziani E, Pomati S, Ramolfo P, et al. Evaluation of retinal nerve fiber layer and ganglion cell layer thickness in Alzheimer’s disease using spectral-domain optical coherence tomography. Invest Ophthalmol Vis Sci. 2013;54(9):5953–5958.

28. Cheung CY, Ong YT, Hilal S, et al. Retinal ganglion cell analysis using high-definition optical coherence tomography in patients with mild cognitive impairment and Alzheimer’s disease. J Alzheimers Dis. 2015;45(1):45–56.

29. Chung SD, Ho JD, Chen CH, Lin HC, Tsai MC, Sheu JJ. Dementia is associated with open-angle glaucoma: a population-based study. Eye (Lond). 2015;29(10):1340–1346.

30. Su CW, Lin CC, Kao CH, Chen HY. Association Between Glaucoma and the Risk of Dementia. Medicine (Baltimore). 2016;95(7):e2833.

31. Lin IC, Wang YH, Wang TJ, et al. Glaucoma, Alzheimer’s disease, and Parkinson’s disease: an 8-year population-based follow-up study. PLoS One. 2014;9(9):e108938.

32. Bach-Holm D, Kessing SV, Mogensen U, Forman JL, Andersen PK, Kessing LV. Normal tension glaucoma and Alzheimer disease: comorbidity? Acta Ophthalmol. 2012;90(7):683–685.

33. Ou Y, Grossman DS, Lee PP, Sloan FA. Glaucoma, Alzheimer disease and other dementia: a longitudinal analysis. Ophthalmic Epidemiol. 2012;19(5):285–292.

34. Kessing LV, Lopez AG, Andersen PK, Kessing SV. No increased risk of developing Alzheimer disease in patients with glaucoma. J Glaucoma. 2007;16(1):47–51.

35. Keenan TD, Goldacre R, Goldacre MJ. Associations between primary open angle glaucoma, Alzheimer’s disease and vascular dementia: record linkage study. Br J Ophthalmol. 2015;99(4):524–527.

36. Harrabi H, Kergoat MJ, Rousseau J, et al. Age-related eye disease and cognitive function. Invest Ophthalmol Vis Sci. 2015;56(2):1217–1221.

37. Jefferis JM, Taylor JP, Collerton J, et al. The association between diagnosed glaucoma and cataract and cognitive performance in very old people: cross-sectional findings from the newcastle 85+ study. Ophthalmic Epidemiol. 2013;20(2):82–88.

38. Yochim BP, Mueller AE, Kane KD, Kahook MY. Prevalence of cognitive impairment, depression, and anxiety symptoms among older adults with glaucoma. J Glaucoma. 2012;21(4):250–254.

39. Weintraub S, Wicklund AH, Salmon DP. The neuropsychological profile of Alzheimer disease. Cold Spring Harb Perspect Med. 2012;2(4):a006171.

40. Kramer JH. Special series introduction: NIH EXAMINER and the assessment of executive functioning. J Int Neuropsychol Soc. 2014;20(1):8–10.

41. Possin KL, LaMarre AK, Wood KA, Mungas DM, Kramer JH. Ecological validity and neuroanatomical correlates of the NIH EXAMINER executive composite score. J Int Neuropsychol Soc. 2014;20(1):20–28.

42. Baldo JV, Delis D, Kramer J, Shimamura AP. Memory performance on the California Verbal Learning Test-II: findings from patients with focal frontal lesions. J Int Neuropsychol Soc. 2002;8(4):539–546.

43. Delis DC, Wetter SR, Jacobson MW, et al. Recall discriminability: utility of a new CVLT-II measure in the differential diagnosis of dementia. J Int Neuropsychol Soc. 2005;11(6):708–715.

44. Donders J. A confirmatory factor analysis of the California Verbal Learning Test--Second Edition (CVLT-II) in the standardization sample. Assessment. 2008;15(2):123–131.

45. Fellman RL, Mattox CG, Ross KM, Specialist AC, Vicchrilli S, Executive AC. Know the New Glaucoma Staging Codes.

46. Kramer JH, Mungas D, Possin KL, et al. NIH EXAMINER: conceptualization and development of an executive function battery. J Int Neuropsychol Soc. 2014;20(1):11–19.

47. Charlson ME, Pompei P, Ales KL, MacKenzie CR. A new method of classifying prognostic comorbidity in longitudinal studies: development and validation. J Chronic Dis. 1987;40(5):373–383.

48. McCoskey M, Addis V, Goodyear K, et al. Association between Primary Open-Angle Glaucoma and Cognitive Impairment as Measured by the Montreal Cognitive Assessment. Neurodegener Dis. 2018;18(5-6):315–322.

